# Evaluation of *Apolipoprotein e4* allele as susceptible factor for neurodegenerative diseases among Eastern Indians

**DOI:** 10.1101/2023.06.21.23291697

**Authors:** Dipanwita Sadhukhan, Smriti Mishra, Priyanka Mukherjee, Atanu Biswas, Subhra Prakash Hui, Tapas Kumar Banerjee, Arindam Biswas

## Abstract

**Introduction:** Apolipoprotein E (ApoE) and age have been identified as the major risk factors for several neurodegenerative disorders. Among the three major isoforms (*ApoE2, ApoE3 & ApoE4*) of *APOE, ApoE4* often shows ethnicity dependent association with neurodegenerative diseases. In the present study, we aim to determine the *e4* allele/genotype frequency among the different neurodegenerative disorders and their correlation with several demographic and clinical parameters from eastern India.

**Material & Methods:** A total of 826 individuals were recruited for this study which includes 128 PD-MCI, 144 PDD, 90 DLB, 114 FTD, 94 AD and 256 unrelated neurologically controls from eastern India. Subjects were analysed for *APOE* genotype (*E2, E3* and *E4*) by PCR-RFLP techniques and Sanger sequencing.

**Results:** The *APOE4* allele was significantly associated with each of the disease subtypes, selected for the study (P = 0.016 to 0.000), while *e3* allele was predominant among controls. Further stratification of subjects identified significant overrepresentation of (a) positive family history for FTD (P = 0.0414) & AD (P = 0.029) and (b) early age at onset of for PDD (P = 0.0316) and FTD (P = 0.0034) among the *e4* allele carriers. Furthermore, lowering of BMSE score was also observed among the *e4* allele carriers of AD subjects (P = 0.0324).

**Conclusion:** There is a significant association of *APOE4* allele with different neurodegenerative diseases influencing lowering of age at onset, BMSE score and positive family history among ethnic Bengali population of eastern India.

## Introduction

The Apolipoprotein E (ApoE) protein has a crucial role in lipid and cholesterol flux in the central nervous system [1]. Among its three major human isoforms, ApoE2, ApoE3, and ApoE4, *APOE4* is considered as one of the most potent risk factors for the development of neurological disorders. The presence of two arginine at the 112^th^ and 158^th^ positions in the APOE4 protein provides it with an enhanced lipid binding property with reduced stability rendering pathogeniecity than other isoforms.

Neuronal death in neurodegenerative diseases is often associated with abnormal protein accumulation, immune system activation, neuroinflammation and blood-brain barrier disruption. In Alzheimer’s disease (AD) pathology, APOE polymorphisms with the combined effect of sex, age, diet, and physical exercise, have a major effect on astrocytic and microglial function and microglial dynamics, synaptic function, amyloid-β load, tau pathology, autophagy, and cell–cell communication [2]. As a result, APOE4 carriers often show a greater Aβ deposition both in quantity and density compared to non-APOE4 carriers. Similarly, in Parkinson’s disease (PD), an increase in alpha-synuclein aggregate formation is found to be influenced by the APOE4 genotype [3]. However, studies showed incongruities in this regard suggesting that APOE polymorphisms may result in a higher risk of cognitive impairment than motor deterioration. In Lewy body dementia, it also has distinct roles in microglia-associated alpha-synuclein clearance in the midbrain [4]. A number of genetic association studies also highlighted increased disease risk for *e4* alleles in frontotemporal dementia [5].

Although APOE plays a significant role in several central nervous system pathologies which are mostly complex in nature, the genetic association data often appear to be inconsistent due to differences in APOE allele frequencies across ethnic populations. In India, a number of genetic studies have been performed in neurodegenerative disorders considering either one or few disease types at a time. However, the results are conflicting and may be influenced by with limited number of study subjects and mutiethnicity. Therefore, the present study aims to evaluate the frequency of APOE isoforms in several neurodegenerative diseases in a relatively larger sample representing the ethnic Bengali population of Eastern India.

## Materials and methods

### Study subjects

A total of 570 patients clinically diagnosed with neurodegenerative disorders [128 Parkinson’s disease with mild cognitive impairment (PD-MCI), 144 Parkinson’s disease with dementia (PDD), 90 Dementia with Lewy body (DLB), 114 Frontotemporal dementia (FTD) and 94 Alzheimer’s disease(AD)] by clinicians were recruited from Eastern India in this study with their written informed consent as per guidelines of Indian Council of Medical Research (ICMR). Patients were diagnosed following standard diagnosis criteria. In addition, 256 age and ethnicity matched unrelated healthy individuals with no personal or family history of neurodegenerative diseases and/ or any other neurological symptoms were also recruited as controls in the present study from Kolkata. Bengali version of Mini Mental State Examination (BMSE) scale was used for primary tool during evaluation [6]. The BMSE score of the recruited control individuals were ≥ 26. The diagnostic labelling was done by trained neurologists (AB and TKB). Diagnosis was performed using standard diagnostic criteria (MDS task force criteria for PD-MCI [7] and PDD [8]), FTD by FTD Consortium criteria [9], DLB by DLB consortium criterion – fourth consensus [10] and AD by NIA-AA criteria [11]. The demographic details of patients are described in Table 1.

**Table 1:** Demographic details of studied population.

| **Category** | **Male:**<br>**Female** | **Familial:**<br>**Sporadic** | **Early onset:**<br>**Late onset** | **Age at onset ±**<br>**SD** | **BMSE ± SD** |
| --- | --- | --- | --- | --- | --- |
| **PD – MCI** (n = 128) | 90: 38 | 32: 96 | 38: 90 | 55.54 ± 15.43 | 21.95 ± 6.13 |
| **PDD** (n = 144) | 100: 44 | 32: 112 | 26: 118 | 60.89 ± 12.94 | 20.23 ± 7.39 |
| **DLB** (n = 90) | 64: 26 | 24: 66 | 16: 74 | 60.65 ± 10.45 | 15.60 ± 8.98 |
| **FTD** (n = 114) | 76: 38 | 20: 94 | 24: 90 | 55.77 ± 10.41 | 15.15 ± 8.67 |
| **AD** (n = 94) | 52: 42 | 48: 46 | 26: 68 | 58.30 ± 9.84 | 14.04 ± 6.91 |
| **CONTROL**<br>(n = 256) | 174 : 82 | Not applicable | Not applicable | Not applicable | $\geq 26$ |

\*PD - MCI: Parkinson's disease with mild cognitive impairment; PDD: Parkinson's disease with dementia; DLB: Dementia with Lewy body; FTD: Frontotemporal dementia; AD: Alzheimer's disease; SD: Standard deviation; Early onset $\leq 50$ years; Late onset $>50$ years; BMSE: Bengali version of Mini Mental State Examination

### Collection of blood samples and genomic DNA preparation

10 mL of peripheral blood samples were collected in EDTA vials from patients and controls. Genomic DNA was prepared from fresh whole blood by conventional salting out method using sodium perchlorate followed by isopropanol precipitation [12] and dissolved in TE (10 mM Tris− HCl, 0.1 mM EDTA, pH 8.0) and stored at 4°C.

### Polymerase chain reaction (PCR), Restriction fragment length polymorphism (RFLP) analysis

*APOE* genotypes were determined by polymerase chain reaction (PCR) and restriction fragment length polymorphism techniques. The amplicon which was generated using specific primer pair was subjected to enzymatic digestions for 3 hours at 37°C with 1 U of the AflIII (New England Biolabs, Ipswich) and HaeII (New England Biolabs, Ipswich) independently. The digested products were separated on a 6% polyacrylamide gel and analysed further after visualization in a Gel documentation system (BIO-RAD, USA).

### Statistical analysis

Demographic and cognitive parameters were analysed using Mann Whitney *U* test and Fisher exact test. Frequency of *APOE* alleles among neurodegenerative disorder subgroups was calculated with healthy controls as a reference group, using Fisher’s exact test. Relative Risk was calculated using the MedCalc statistical software. The probability level of ≤ 0.05 was considered statistically significant.

## Results

### Frequency of *APOE4* in disease subgroups

The *APOE* SNPs, rs429358 and rs7412, which encode for the polymorphisms ε2, ε3, and ε4 were screened in 570 patients with various neurodegenerative disorders against 256 ethnically matched controls. All APOE alleles were within Hardy-Weinberg equilibrium in both the patients and control groups. Table 2 shows the allele and genotype frequencies of APOE variants in different disease types. For the *APOE4* allele, a significant association with PD-MCI, PDD, DLB, FTD and AD were observed in our study cohorts (P = 0.016 for PD-MCI and 0.000 for rest). The highest relative risk was observed for the AD group (RR = 5.59; 95% CI: 4.44 – 7.60; P = <0.0001) followed by DLB (RR = 3.89), FTD (RR = 2.54), PDD (RR = 2.53) and PD-MCI (RR = 1.79) [Table 3]. In contrast, the e3 allele was the predominant one in control group.

**Table 2:**
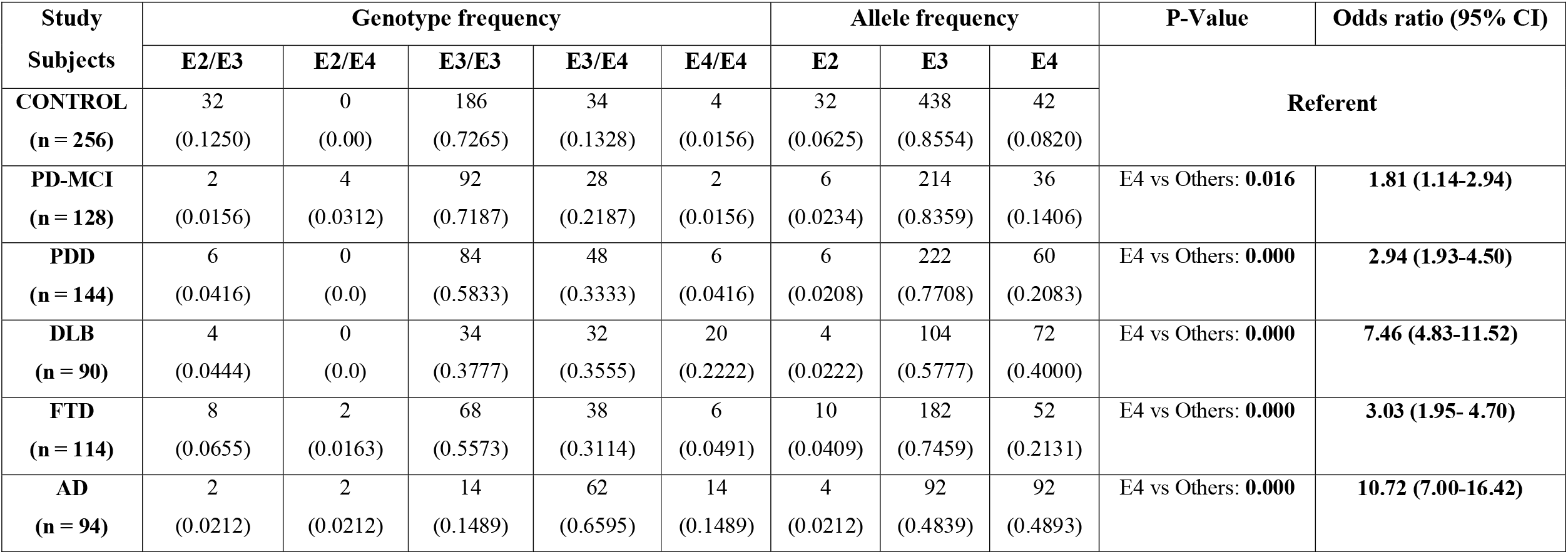
Allele & Genotype frequency of *APOE* among different neurodegenerative diseases.

**Table 3:** Frequency of APOE4 carriers among different neurodegenerative disorders.

| **Diagnosis** | **Subjects**<br>**(n)** | **E4 Carriers**<br>**(n)** | **% of**<br>**Subjects** | **\*\*P value** | **Relative Risk (95% CI)** | **P value** |
| --- | --- | --- | --- | --- | --- | --- |
| **PD-MCI** | 128 | 34 | 26.56% | 0.0081 | 1.79 (1.186 – 2.699) | 0.0055 |
| **PDD** | 144 | 54 | 37.5% | < 0.00001. | 2.53 (1.760 – 3.626) | <0.0001 |
| **DLB** | 90 | 52 | 57.78% | < 0.00001. | 3.89 (2.763 – 5.482) | <0.0001 |
| **AD** | 94 | 78 | 82.98% | < 0.00001. | 5.59 (4.110 – 7.602) | <0.0001 |
| **FTD** | 122 | 46 | 37.70% | < 0.00001. | 2.54 (1.752 – 3.683) | <0.0001 |
| **Controls** | 256 | 38 | 14.84% | References | References | References |

\*\*Fisher exact test was calculated considering healthy controls as reference group; Relative risk was calculated using MedCalc software.

### Comparison of demographic features and cognitive status

Next, we compared the demographic features like age at disease onset, positive family history, and gender between carriers and non-carriers of *APOE E4* variant in each of the disease subtype. A significant lowering of age at onset for PDD (56.85 years vs 63.08 years; P = 0.0316) and FTD (54.09 years vs 58.79 years; P = 0.0034) groups was observed for *E4* carriers, while the presence of positive family history for FTD (P = 0.0414) and AD cases (P = 0.029) show significant correlation for the same [Table 4].

**Table 4:**
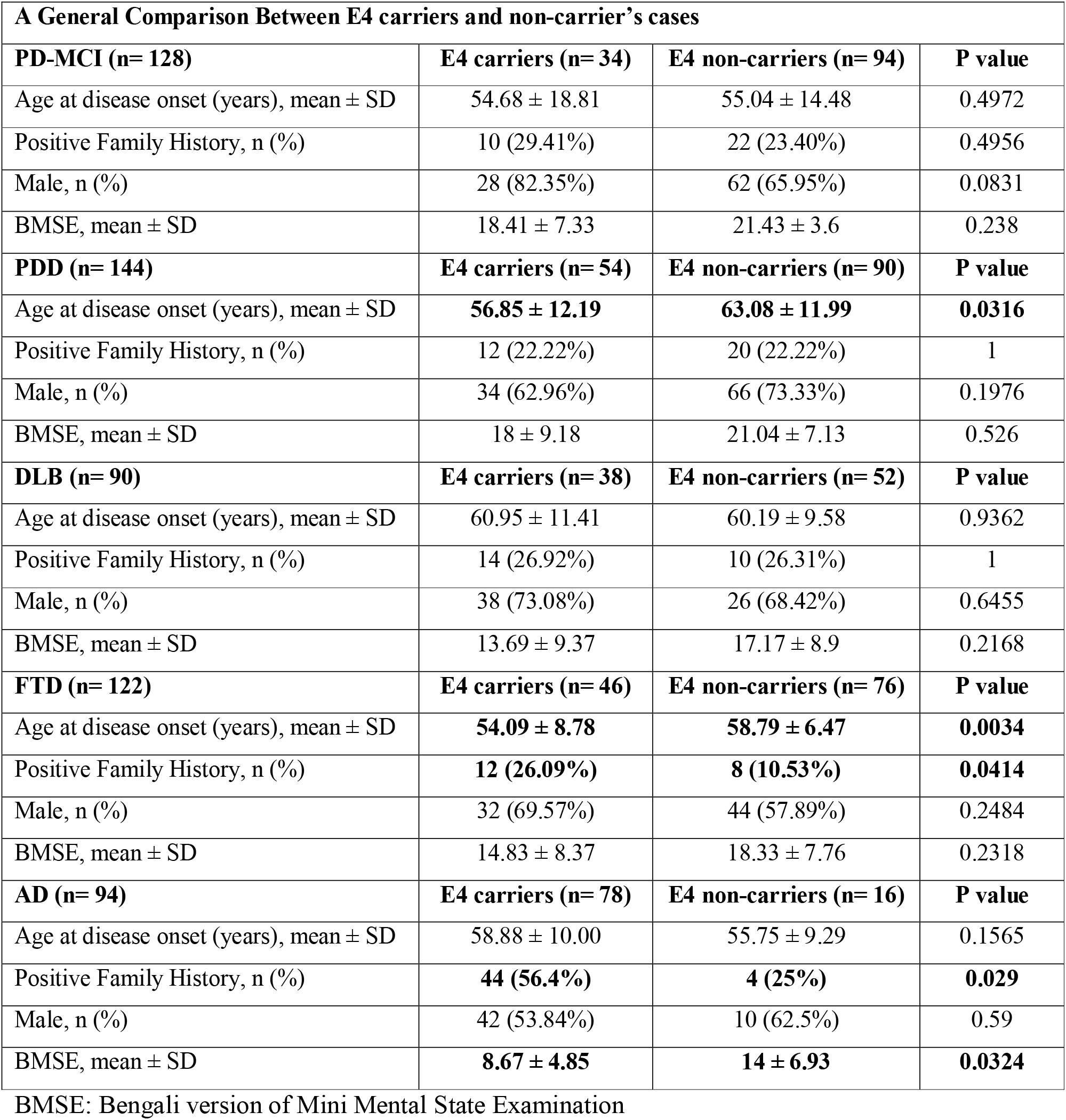
Comparison of clinical parameters between E4 carriers and E4 noncarriers.

As mentioned in Table 1, irrespective of *APOE4* status a lowering of BMSE score *i*.*e* <25 (referring overall cognitive status) was observed in our patient cohort with different clinical diagnosis. However, after further stratification of subjects according to E4 isoform followed by an intra-group comparison, a significant lowering in BMSE score for AD (P = 0.0324) but not for the other conditions was observed between E4 carriers and non-carriers.

## Discussion

Identification of population-specific genetic risk variants in complex neurodegenerative diseases has long been reported in different studies. However, the major gene of interest is APOE across the world population showing ethnicity and disease specific association. In this present study, we observed an overrepresentation of APOE e4 allele in AD, PD, PD-MCI, DLB and FTD cases in ethnic Bengali population. Our data corroborates with previous Indian reports on PD, MCI and AD but is at variance with the other studies on DLB, FTD and FTLD [Table 5].

**Table 5:**
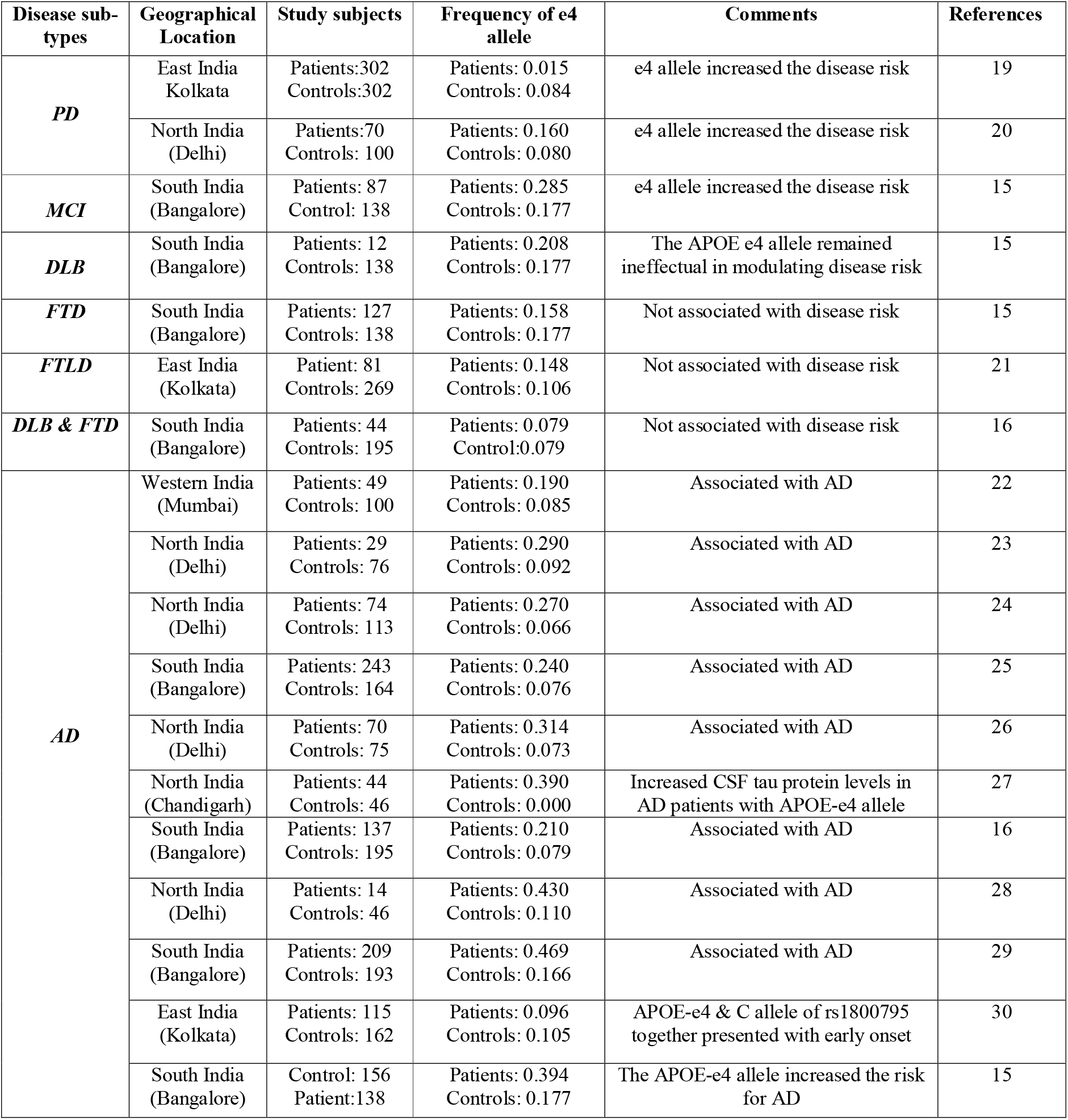
Frequency distribution of APOE4 allele as reported in Indian studies on neurodegeneration.

AD and PD cases are often been categorized into early-onset or late-onset cases, as age is an independent risk factor for both. Recent evidences suggest that although the APOE2 allele decreases late onset AD (LOAD) risk, the expression of one and two E4 allele increases the disease risk to threefold and 9 to 15-fold, respectively [13]. However, no such findings were observed here which may be due to recruitment of only late-onset AD cases and small number of non-E4 carrying AD patients in study cohort as well. In contrast, APOE4 allele carriers showed an early age at onset for FTD and PDD cohort [5]. Our present data for PDD is consistent with a meta-analysis study in 2006 [14]. However, negative association was reported in an early report from South India on FTD [15, 16]. The small number of samples and genetic divergence may be probable reasons for lack of consistency in the nature of association between two studies from India. The south Indian population is representative of the original Dravidians from Central and South India while Eastern Indians belong to the Indo-European (IE) lineage [17].

Evidences suggest that positive family history in neurodegenerative disorders explains the part of individual differences in risk and progression for the diseases and thus it is often correlated with genetic variants to determine its influence in disease pathology. Here, an intra group comparative study describing the correlation between APOE4 polymorphism and family history in different diseases as summarised in Table 4, revealed a link only for AD and FTD suggesting its potential for these diseases among the Eastern Indians. However, the absence of such in rest suggests the role of additional genes with equal or greater significance.

The progression of neurodegeneration is always accompanied by different degrees of cognitive decline. Diseases like AD, DLB, FTD manifest cognitive failure as an initial symptom while PD-MCI and PDD are developed in few PD patients over time. A number of earlier reports, on the basis of frequency differences between cases and controls, already stated the E4 allele to be correlated with higher prevalence of dementia in synuclenopathies and AD [13, 18]. However, when we compared the raw scores according to genotype, except for AD, no other diseases showed differences in BMSE scores between carriers and non-carriers among the patient cohort. Our data is concordant with other national and international reports for AD, while it needs further confirmatory studies considering scores and further sub domain analysis to make a definite conclusion.

## Conclusion

In conclusion, our findings demonstrate that the APOE4 allele increases the risk of AD, PDD, PD-MCI, DLB and FTD among ethnic Bengali populations from Eastern India. Additionally, it has a negative impact on cognitive status for AD while influencing the early age at onset for PDD and FTD. A positive correlation between family history and APOE4 was also identified for AD and FTD. It would also be interesting to study and correlate the impaired cognitive subdomains in different disease types with APOE genotypes in future studies.

## Data Availability

The data described in this study are available from the corresponding author upon reasonable request.

## Ethics statement

All procedures performed in studies involving human participants were in accordance with the ethical standards of the institutional and national research committee as well as the 1964 Helsinki Declaration and its later amendments.

## Informed consent

Informed consent from all the participants were received prior to clinical data and sample collection.

## Acknowledgement

The authors thank the patient, family members, and healthy individuals who participated in the study.

## Funding

Supported by grants from the Department of Science & Technology, Govt. of India, under Cognitive Science Research Initiative Programme (DST/CSRI-PDF/2021/12) and (SR/CSRI/PDF-32/2014 and DST/CSRI-P/2017/22).

## Author’s contribution

Dipanwita Sadhukhan and Arindam Biswas were responsible for the concept, study design and manuscript preparation. Atanu Biswas and Tapas Kumar Banerjee were responsible for the diagnosis, recruitment, clinical evaluation and blood collection for all study subjects. Smriti Mishra, Priyanka Mukherjee were responsible for experimental work and manuscript preparation. Subhra Prakash Hui was responsible for manuscript preparation. All authors read the draft, provided their inputs and agreed on the final version of the manuscript.

## Conflict of Interest

The authors declare no conflicts of interest.

## Notes

### Competing Interest Statement

The authors have declared no competing interest.

### Author Declarations

IPGME&R RESEARCH OVERSIGHT COMMITTEE AND APPROVED

